# Prediction models with survival data: a comparison between machine learning and the Cox proportional hazards model

**DOI:** 10.1101/2022.03.29.22273112

**Authors:** Audinga-Dea Hazewinkel, Hans Gelderblom, Marta Fiocco

## Abstract

Recent years have seen increased interest in using machine learning (ML) methods for survival prediction, chiefly using big datasets with mixed datatypes and/or many predictors Model comparisons have frequently been limited to performance measure evaluation, with the chosen measure often suboptimal for assessing survival predictive performance. We investigated ML model performance in an application to osteosarcoma data from the EURAMOS-1 clinical trial (NCT00134030). We compared the performance of survival neural networks (SNN), random survival forests (RSF) and the Cox proportional hazards model. Three performance measures suitable for assessing survival model predictive performance were considered: the C-index, and the time-dependent Brier and Kullback-Leibler scores. Comparisons were also made on predictor importance and patient-specific survival predictions. Additionally, the effect of ML model hyper-parameters on performance was investigated. All three models had comparable performance as assessed by the C-index and Brier and Kullback-Leibler scores, with the Cox model and SNN also comparable in terms of relative predictor importance and patient-specific survival predictions. RSFs showed a tendency for according less importance to predictors with uneven class distributions and predicting clustered survival curves, the latter a result of tuning hyperparameters that influence forest shape through restrictions on terminal node size and tree depth. SNNs were comparatively more sensitive to hyperparameter misspecification, with decreased regularization resulting in inconsistent predicted survival probabilities. We caution against using RSF for predicting patient-specific survival, as standard model tuning practices may result in aggregated predictions, which is not reflected in performance measure values, and recommend performing multiple reruns of SNNs to verify prediction consistency.

## 1 Introduction

Over the last decade, interest and publications on machine learning approaches in medical research and, specifically, cancer research have grown, resulting in a still ongoing debate on the value of machine learning (ML) approaches versus more traditional statistical modelling (SM). Statistical models exist within a mathematical framework, make probabilistic assumptions about the data generation process, and generally yield easily interpretable results. In contrast, ML approaches do not impose an *a priori* specified relationship on the predictors and outcome, and rely on a data-driven, algorithmic approach. While the absence of preconceived structures allows for complicated non-linear effects and high-order interactions between variables, these relationships cannot be accurately quantified, and, at best, only a general indication of predictor importance can be obtained (Kourou *et al*., 2015; Shahid *et al*, 2019; Sidey-Gibbons & Sidey Gibbons, 2019). In this article, we investigate the predictive performance of neural networks (Bishop, 2006; Hertz *et al*., 1991; Minsky & Papert, 1969) and random forests (Breiman, 2001) in an application to time-to-event data, and compare it to the performance of the Cox proportional hazards model (Cox, 1972), which is traditionally the analysis model of choice for clinical survival data. Previous investigations of the potential of ML methods have chiefly focused on big datasets with large numbers of predictors – for example genomic or microarray data. Here, we consider the effect of seven variables on overall survival, three of which have previously been shown to be strongly associated with decreased survival. Our choice of data has a two-fold purpose. In addition to assessing the potential of ML for clinical data with smaller numbers of predictors, the comparatively simple data structure and *a priori* available clinical knowledge permits us to establish a reasonable expectation of results, allowing us to identify ML pitfalls that would remain hidden in more complex data types. We apply these methods to osteosarcoma patients from the EURAMOS-1 clinical trial (NCT00134030) (Whelan *et al*., 2012). Osteosarcoma is the second most common primary bone tumour, and is primarily diagnosed in adolescents and young adults (Bacci *et al*., 1990; Bielack *et al*., 2002; Rosen *et al*., 1976; Whelan *et al*., 2012).

## 2. Methods

### 2.1 Artificial neural networks for survival data

Artificial neural networks (ANNs) rely on nodes (neurons) arranged in a structure that consists of an input layer that contains the predictor variables, at least one hidden layer, and an output layer with one or more nodes representing the outcome, with the nodes in a given layer connected to each nodes in the subsequent one (Bishop, 2006; Hertz *et al*., 1991; Minsky & Papert, 1969). The number of hidden layers and corresponding nodes are user-specified, with greater numbers allowing for more flexible and complex relationships between the variables. The amount of information passed from the input layer to the output layer is modulated by transfer functions. Weights are estimated for the connections between nodes and are used to predict the outcome. The model is fitted by minimizing an error function in an algorithmic approach, which, if correctly specified, will ensure the difference between observed and predicted values is also minimized. To prevent a model from becoming too attuned to the training data, and consequently generalizing poorly to new data, regularization can be imposed by means of a weight decay parameter (Shahid *et al*, 2019; Minsky & Papert, 1969).

Traditionally, ANNs are used to classify subjects into two or more classes. Biganzoli *et al*. (1998) and Kantidakis *et al*. (2020) adapted ANNs to time-to-event data, by redefining the survival problem as a classification problem and dividing the follow-up time into intervals. To estimate the survival neural network (SNN), the data is transformed longitudinally, with each patient replicated for the number of observed intervals and a binary event indicator denoting whether an event occurred in each interval. This allows a straightforward neural network to be fitted, with an additional interval variable that, for each entry, denotes the corresponding time interval. Biganzoli shows that by minimizing a cross-entropy error function and specifying logistic transfer functions the likelihood is maximized and the conditional death probability (the hazard) is estimated. From the latter, survival probabilities can be easily obtained.

### 2.2 Random survival forests

Random forests (RFs) consist of decision trees, which are grown from many bootstrap samples from the original data. Decision trees rely on binary recursive partitioning of a given predictor space into increasingly smaller and homogeneous regions until a certain stopping criterion is reached. These regions are referred to as nodes, with the final regions of a tree called terminal nodes. Each node split is determined by which combination of predictor and predictor splitting point results in the biggest difference between the two resulting daughter nodes (Breiman, 2001; Strobl *et al*., 2009; Zhou & McArdle, 2015). Ishwaran *et al*. (2008) developed the random survival forest (RSF) approach, where the increase of homogeneity is achieved by maximizing the survival difference, as measured by the log-rank test. The trees are grown under the constraint that at least one unique event be present in each terminal node. The cumulative hazard is calculated for each terminal node using the Nelson-Aalen estimator. To obtain the cumulative hazard, and by extension the survival function, for a new subject, the predictor vector is ‘dropped’ down the tree, into a single unique terminal node. As single decision trees are unstable and sensitive to minor changes in the data, trees are grown from many bootstrap samples, and the results averaged – a procedure known as bagging. RFs diverge from bagging by introducing an additional source of variability and selecting a random subset of predictors as candidates for a node split (Breiman, 2001; Ishwaran *et al*. 2008; Ishwaran & Kogalur, 2008; Strobl *et al*., 2009; Zhou & McArdle, 2015).

### 2.3 Model tuning

Both ANNs and RFs use hyperparameters that should be tuned in a cross-validation approach. For ANNs, these constitute the number of nodes in the hidden layer and the weight decay parameter value (Bishop, 2006; Hertz *et al*., 1991; Minsky & Papert, 1969). For RSFs the hyperparameters are: 1) *mtry*: number of candidate variables considered at each split; 2) *nsplit*: number of randomly selected splitting points on each variable; 3) *nodesize*: average number of observations per terminal node; 4) *nodedepth*: maximum number of node splits from root to terminal node. *Nodesize* and *nodedepth* allow for the most explicit regulation of tree growth, which can be restricted by specifying large and small values, respectively (Ishwaran *et al*. 2008; Ishwaran & Kogalur, 2008). To tune and train the models, the data were divided into a training (67%) and validation set (33%), while maintaining the event/censoring proportion of the original data. The hyperparameters were tuned in a 5-fold cross-validation, performed on the training set. For the SNN, the 8.9 year follow-up time was divided in 107 1-month long intervals, and the tuning process used to find the best combination of 1 to 10 hidden nodes and weight decay parameter values of 0.0001, 0.001, 0.01, 0.05 and 0.1. For the RSF, the four parameters were tuned in a grid approach, considering 1 to 7 candidate splitting variables (*mtry*), node sizes from 1 to 400 in increments of 20 (*nodesize)*, 1 to 10 split points *(nsplit*), and tree depths of 1 to 15 (*nodedepth*). Each forest was comprised of 750 bootstrapped decision trees. Model performance while tuning was assessed with the C-index, with final model performance assessed on the validation set, using additional measures.

### 2.4 Assessing model performance

Model performance was assessed using three measures commonly used for survival data. For the Cox model, SNN and RSF, the Brier score (Graf *et al*. 1999; Van Houwelingen & Le Cessie, 1990; Van Houwelingen & Putter, 2012) and Kullback-Leibler (KL) score (Van Houwelingen & Putter, 2012) were obtained, which use the predicted survival probabilities and the observed event status to quantify the prediction error of a survival model for a given time-point. Score values range from 0 to 0.25 and 0 to 0.69 for the two measures, respectively, with each upper limit given by the value that would be calculated under a null model, and low values indicating good model performance. Discriminative ability was assessed with Harrell’s concordance index (C-index), which calculates the proportion of observations pairs for which the model predictions and observed survival times are concordant (Harrel *et al*. 1996). Values range from 0.5 to 1, with a C-index of 0.5 indicating a model with no discriminative ability and a C-index of 1 a perfect model. The C-index was obtained using the linear prognostic index (PI) and the predicted ensemble mortality, for the Cox and RSF models, respectively. The flexibility of ANNs precludes a natural ordering of subjects, making it impossible to directly obtain a PI-like measure. Instead, we computed a non-linear time-dependent PI for each individual, given by the log-odds ratio of the conditional hazard probabilities predicted by the SNN model (Antolini *et al*., 2005), which were then averaged across the time intervals to obtain an overall PI.

### 2.5 Assessing variable importance

For the Cox model, the effect of a predictor on the outcome is given by a coefficient or hazard ratio. To quantify the relative predictor importance, we calculated Heller’s relative explained risk (Heller, 2014). For ANNs, the weights of the converged neural network can be used to calculate a measure of variable importance by applying Olden and Jackson’s (2002) connection weight method, which sums the products of the weights connecting the relevant input node (predictor) to the hidden layer, and the weights connecting the hidden nodes to the output. For RSFs, the variable contribution is quantified through the variable importance (VIMP) measure, which is given by the difference in prediction error, calculated as 1-C-index, between an RSF fitted to the observed data and data permuted in the variable of interest (Ishwaran *et al*. 2008; Ishwaran & Kogalur, 2008).

### 2.6 Implementation

All analyses were performed in `R`, version 4.0.2 (R Foundation for Statistical Computing). For the RSFs, the package `randomForestSRC` was used (Ishwaran & Kogalur, 2018). For the SNN, no R implementation is currently available. Given the appropriate manual data transformation, the package nnet (Ripley, 2016) can be used. Heller’s relative risk was calculated using the `coxphERR` function from the `clinfun` package (Sheshan, 2018). Missing values were imputed in a 10-fold imputation approach, using the `Amelia` package (Honaker *et al*., 2011). Supplementary R Code is provided for replicating the results on simulated data.

## 3. Data

The EURAMOS (European and American Osteosarcoma Studies) collaboration recruited a total 2260 patients to the EURAMOS-1 trial, from 2005 to 2011. Patients received neadjuvant chemotherapy prior to resection of the primary tumour, after which 1336 patients were randomised to treatment. Patients with a poor histological response (≥ 10% viable tumour), as assessed in the resected specimen, received MAP or MAP with ifosfamide and etoposide, and patients with a good response (< 10%) were allocated MAP or MAP with pegylated interferon (Whelan *et al*., 2012). As no benefit of experimental treatment was found in the primary analysis, we considered all 2260 patients, regardless of randomisation status. We investigated the effect of seven predictors (Table 1) on overall survival, measured in years since surgery, for 2032 patients, excluding those who had no surgery date or follow-up, and randomised patients who were later found ineligible per the study protocol, due to presence of an unresectable disease or the progression or development of a new metastatic disease. (Figure 1). Table 1 shows the patient distribution across predictor categories. No major differences were observed between randomised and non-randomised patients. A good histological response and the presence of metastases have previously been associated with improved and poor survival, respectively adults (Bacci *et al*., 1990; Bielack *et al*., 2002; Rosen *et al*., 1976; Whelan *et al*., 2012).

**Table 1.** Patient demographics and disease characteristics

| Predictor | Randomised |  | Not randomised |  | Total |  |
| --- | --- | --- | --- | --- | --- | --- |
|  | N/mean(SD) | % | N/mean(SD) | % | N/mean(SD) | % |
| Age | 15.0 (5.3) | 100 | 15.1 (5.1) | 100 | 15.0 (5.3) | 100 |
| Missing | 0 | 0 | 0 | 0 | 0 | 0 |
| Histological response<sup>a</sup> |  |  |  |  |  |  |
| Good (<10% tumor) | 715 | 54 | 289 | 41 | 1004 | 49 |
| Poor (≥10% tumor) | 609 | 46 | 349 | 50 | 958 | 47 |
| Missing | 4 | 0 | 66 | 9 | 70 | 3 |
| Excision |  |  |  |  |  |  |
| Wide/radical | 1096 | 83 | 551 | 78 | 1647 | 81 |
| Marginal | 175 | 13 | 73 | 10 | 248 | 12 |
| Intralesional/other<sup>b</sup> | 57 | 4 | 80 | 11 | 137 | 7 |
| Missing | 0 | 0 | 0 | 0 | 0 | 0 |
| Volume<sup>c</sup> | 195 (255) | 81 | 215 (289) | 86 | 202 (268) | 83 |
| Missing | 253 | 19 | 102 | 14 | 0 | 17 |
| Sex |  |  |  |  |  |  |
| Female | 546 | 41 | 287 | 41 | 833 | 41 |
| Male | 782 | 59 | 417 | 59 | 1199 | 59 |
| Missing | 0 | 0 | 0 | 0 | 0 | 0 |
| Lung metastases |  |  |  |  |  |  |
| No | 1083 | 82 | 556 | 79 | 1639 | 81 |
| Yes/Possible | 245 | 18 | 148 | 21 | 393 | 19 |
| Missing | 0 | 0 | 0 | 0 | 0 | 0 |
| Other metastases |  |  |  |  |  |  |
| No | 1279 | 96 | 674 | 96 | 1953 | 96 |
| Yes/Possible | 49 | 4 | 30 | 4 | 79 | 4 |
| Missing | 0 | 0 | 0 | 0 | 0 | 0 |

a) A good and poor histological response is defined by the amount of tumour remaining after resection <10% and ≥10% constitute a good and poor response, respectively.
b) A subset of patients had an excision marked as “unknown”. In the analysis this category was combined with intralesional excision.
c) Absolute volume is measured in cm X cm X cm X 0.54 (spheric T vol), or 0.785 for cylindric T vol)

**Figure 1.**
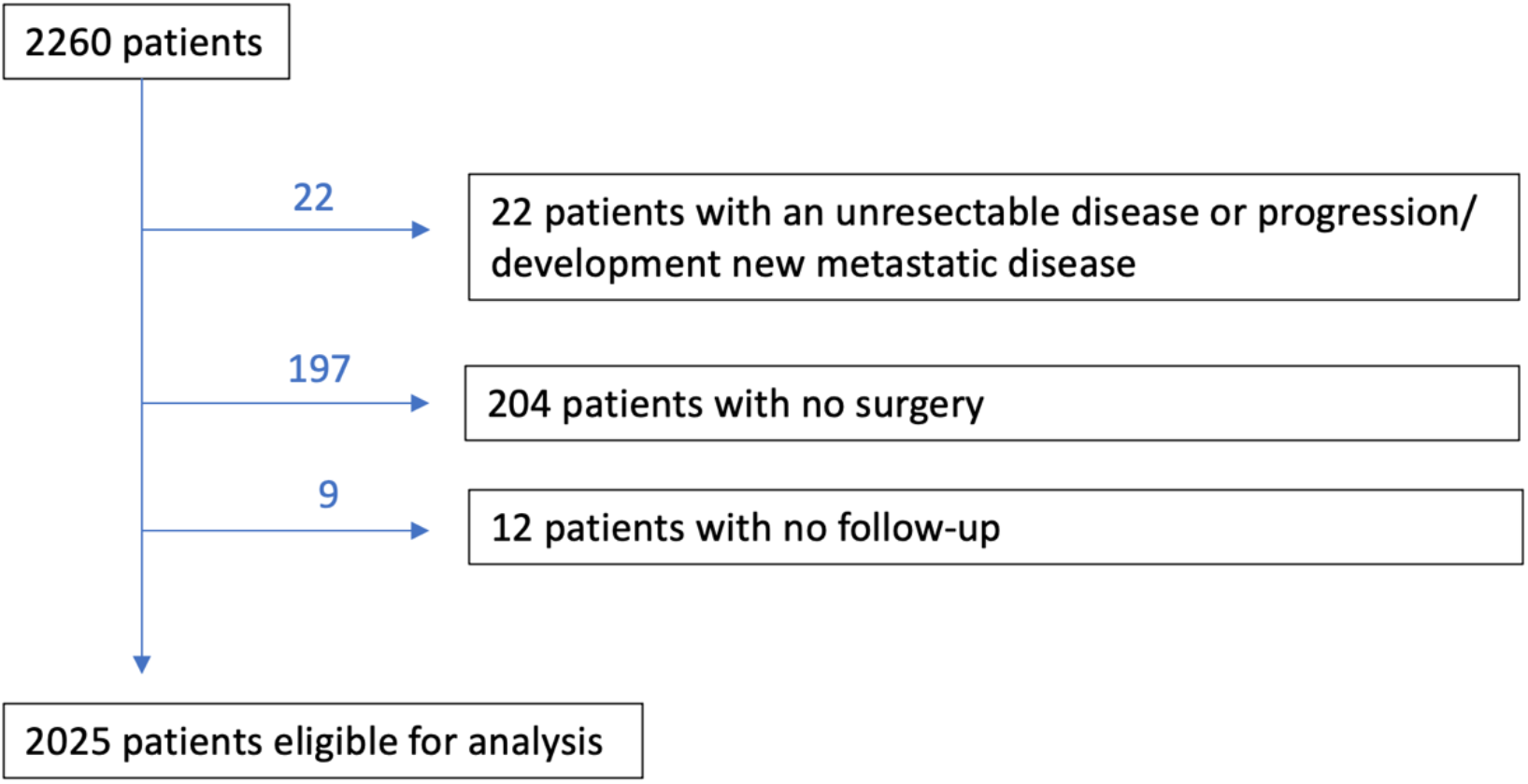
Patient exclusion diagram.

## 4. Results

Maximum follow-up time since surgery was 8.9 years, with a median follow-up time of 4.96 years (95% CI 4.87-5.08), as estimated using the reverse Kaplan-Meier approach (Schemper & Smith, 1996). At end of study, 460 patients had died, while 1572 remained alive. The effect of seven predictors on overall survival is examined in three models. The models are compared on performance as measured by the three performance measures, on predictor importance, and, visually, on pattern and shape of predicted survival curves.

### 4.1 Performance measures

Cross-validation tuning resulted in an SNN with 5 hidden nodes and a decay parameter of 0.05, and an RSF with *mtry*=2, *nsplit*=3, *nodedepth*=9, and *nodesize*=160. Applying these models to the validation data yielded C-indices of 0.707, 0.695, and 0.706 for the Cox, SNN and RSF models, respectively.

Figure 2a shows the time-dependent Brier and KL scores, where lower values indicate better performance. Similar patterns were observed for the Cox model and RSF, with for the SNN moderately lower values in the second half of follow-up. As RSF predictions can only be obtained for event times observed in the training data, no scores were calculated for times greater than 6.9 years after surgery.

**Figure 2.**
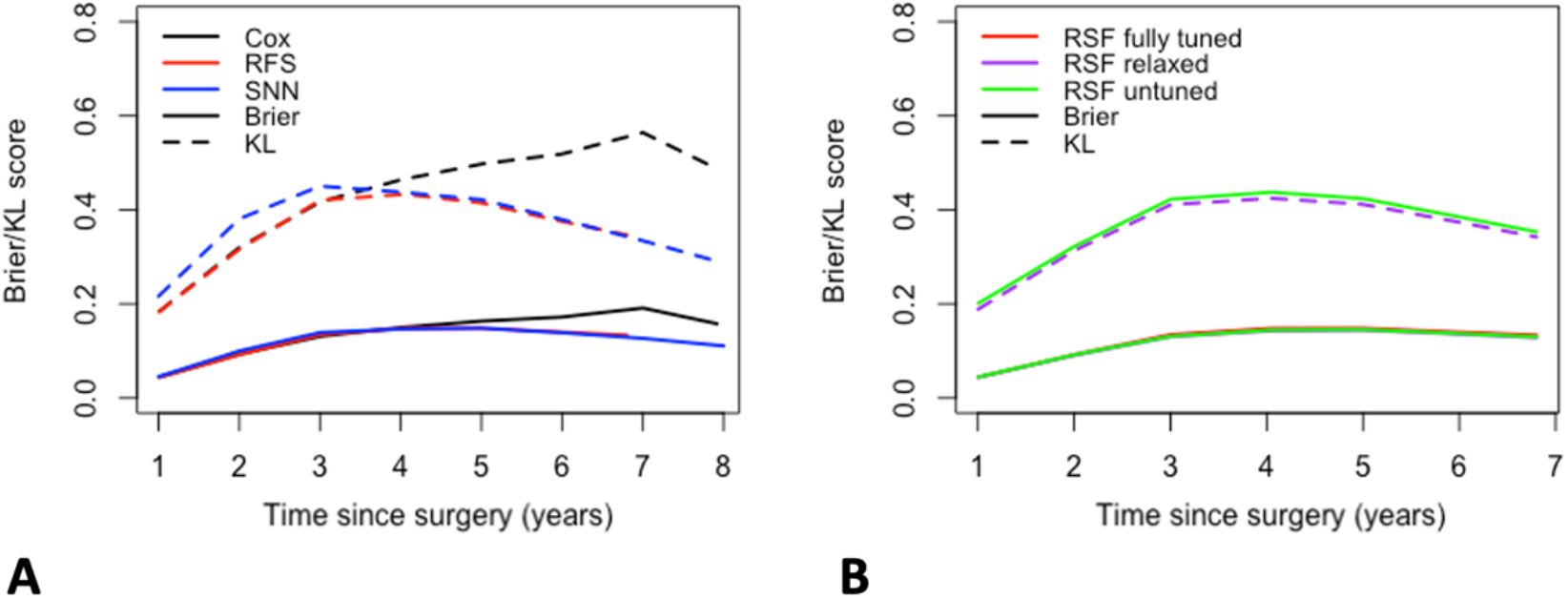
Brier (solid line) and Kullback-Leibler (KL, dashed line) scores for **A)** the Cox proportional hazards model (black), the SNN (blue) and the fully tuned RSF (red); **B)** the fully tuned RSF (red), the RSF with *nodesize* and *nodedepth* restrictions relaxed (purple) and the untuned RSF with no parameter restraints (green).

### 4.2 Mean predicted survival

Figure 3A shows the mean predicted survival probabilities and prediction range for the validation set, with comparable mean probabilities for the Cox (black) and RSF (red) models and lower values for the SNN (blue) at later time points. While both the Cox model and SNN had a comparable range, the RSF predictions showed severe clustering. Model tuning resulted in an RSF with restrictions on both *nodesize* (9) and *nodedepth* (160), limiting the range of values that can be predicted for any new patient. To futher investigate this, we considered an additional two RSF models, one relaxed model (RSF relaxed) with *nodesize* and *nodedepth* unrestricted, and one completely untuned model (RSF untuned), with *nodesize* and *nodedepth* unrestricted, all seven variables selected for each node split (*mtry*=7), and the software default value for the number of random splits considered on each splitting variable (*nsplit*=10). Both models were near identical to the fully tuned RSF, when compared on performance measures, with overlapping Brier and KL scores (Figure 2B) and C-indices of 0.706, 0.694 and 0.677, for the fully tuned, relaxed, and untuned RSF models, respectively. While the mean survival probabilities were also largely unaffected, relaxing *nodesize* and *nodedepth* (purple) resulted in a much wider prediction range, with the widest spread observed when all hyperparameters were relaxed (green, Figure 3B).

**Figure 3.**
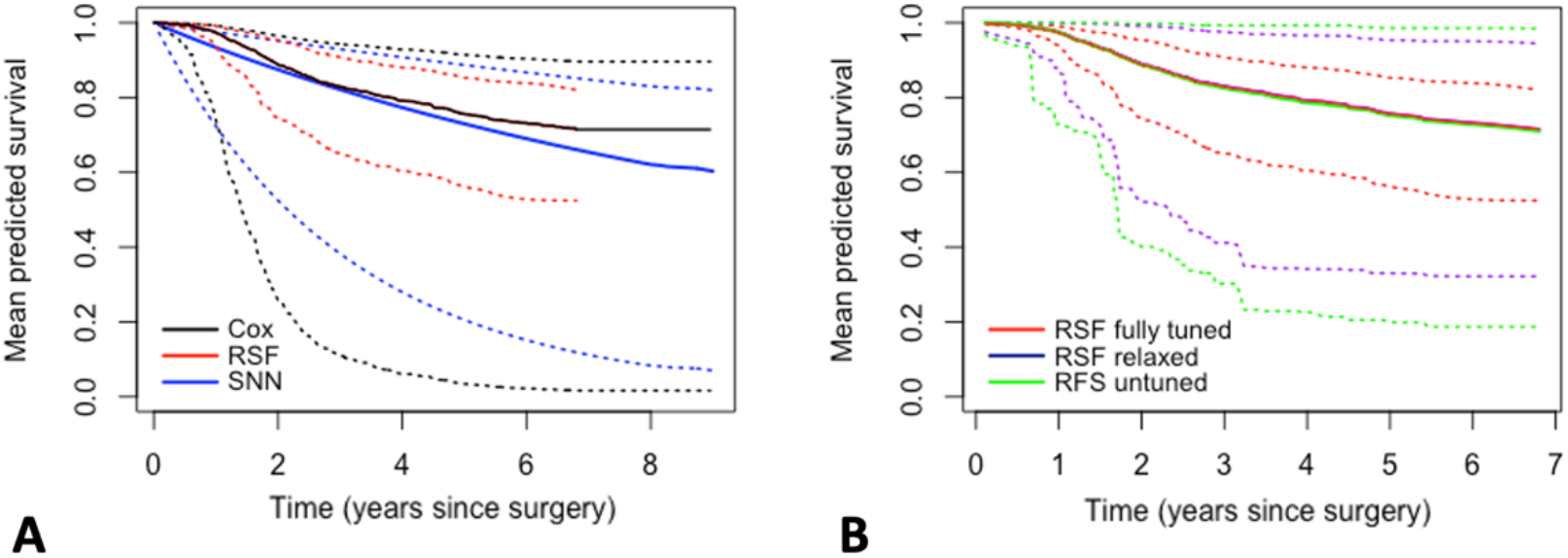
Mean predicted survival probabilities (solid line) and maximum range (dotted line) for **A)** the Cox proportional hazards model (black), the SNN (blue) and the fully RSF (red); **B)** the fully tuned RSF (red), the RSF with *nodesize* and *nodedepth* restrictions relaxed (purple) and the untuned RSF model with no parameter restraints (green).

### 4.3 Variable importance

For the Cox model, we obtained the HRs with 95% CIs for overall survival, and Heller’s explained relative risk (RR), which quantifies the relative variable importance (Table 2). We observed the biggest HRs for the presence of lung metastases, poor histological response, the presence of other metastases, and an intralesional/other excision. Heller’s RR largely maintained this order of importance but identified volume as the third strongest predictor. The SNN results diverged slightly, prioritizing other metastases over histological response and identifying a marginal excision as an important contributor (Figure 4A). For the fully tuned RSF, the order of importance was comparable to the Cox model, with the highest *VIMP* observed for lung metastases, followed by histological response, volume and other metastases (Figure 4B). Relaxing the RSF resulted in a decreased VIMP value for the first three predictors, and an increase in VIMP for volume and excision (Figure 4C), with, for the untuned RSF, volume identified as the most important predictor (Figure 4D).

**Table 2.** Hazard ratio (HR) estimates from the Cox proportional hazards model, with 95% confidence intervals (CIs) and Heller’s relative risk (RR) estimates.

| Predictor | HR | 95% CI | Heller's RR<sup>d</sup> |
| --- | --- | --- | --- |
| Lung metastases |  |  |  |
| No | 1 |  |  |
| Yes/possible | 2.565 | 2.02-3.257 | 0.1028 |
| Histological response<sup>a</sup> |  |  |  |
| Good (<10% tumor) | 1 |  |  |
| Poor (≥10% tumor) | 2.167 | 1.711-2.743 | 0.0922 |
| Other metastases |  |  |  |
| No | 1 |  |  |
| Yes/possible | 2.068 | 1.332-3.213 | 0.0068 |
| Excision |  |  |  |
| Wide/radical | 1 |  |  |
| Marginal | 0.776 | 0.539-1.118 | 0.0041 |
| Intralesional/other<sup>b</sup> | 1.48 | 0.993-2.207 | 0.0068 |
| Sex |  |  |  |
| Female | 1 |  |  |
| Male | 1.148 | 0.908-1.451 | 0.0028 |
| Volume<sup>c</sup> | 1.000 | 1-1.001 | 0.0115 |
| Age | 0.999 | 0.978-1.02 | 0.0000 |

a) A good and poor histological response is defined by the amount of tumour remaining after resection <10% and ≥10% constitute a good and poor response, respectively.
b) A subset of patients had an excision marked as "unknown". In the analysis this category was combined with intralesional excision.
c) Absolute volume is measured in cm X cm X cm X 0.54 (spheric T vol), or 0.785 for cylindric T vol)
d) The contribution of each variable to Heller's explained relative risk was calculated. The relative risk for the full model was 0.275 (SE 0.04).

**Figure 4.**
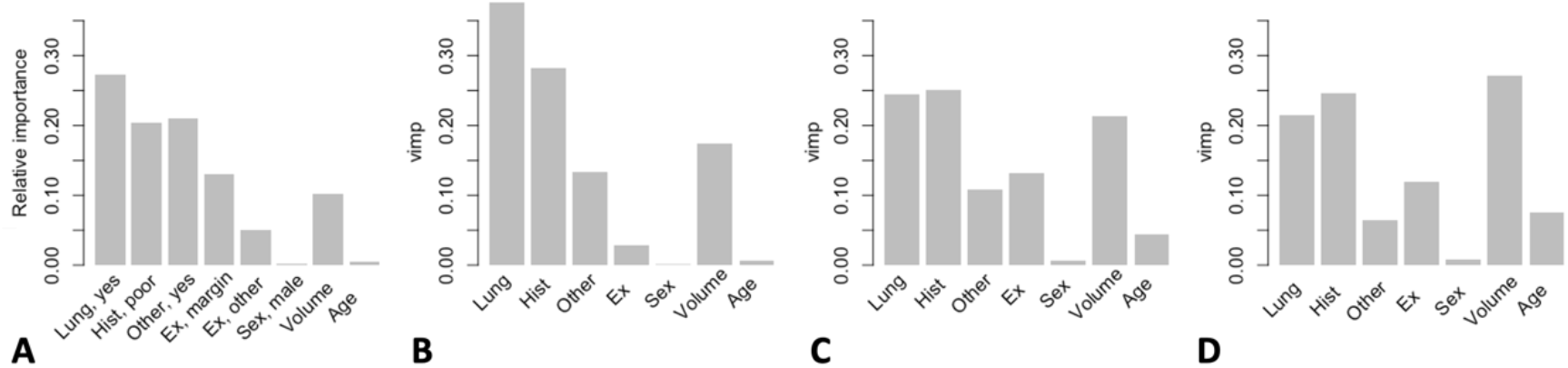
Variable importance for the SNN and RSF models. **A)** Relative variable importance measures for the SNN as calculated using Ogden and Jackson’s connection weight method, with estimates for presence of lung metastases (Lung, yes), poor histological response (Hist, poor), presence of other metastases (Other, yes), marginal excision (Ex, margin), intralesional/other excision (Ex, other), male sex (Sex, male), absolute tumour volume (Volume) and age at surgery (Age). Reference categories are given in Tables 1 and 2; **B)** Relative variable importance measure (VIMP) for the fully tuned RSF; **C)** Relative VIMP for the RSF model with *nodesize* and *nodedepth* restrictions relaxed; **D)** Relative VIMP for the untuned RSF model with all parameters unrestrained.

### 4.4 Predicted survival curves

In Figure 5 the predicted survival curves are plotted for all validation set patients, and we observe the same general pattern as for the mean predicted survival plots (Figure 3), with wide prediction spreads for the Cox (Figure 5A) and SNN (Figure 5B) models, clustered predictions for the fully tuned RSF (Figure 5C), and a wider range for the relaxed (Figure 5D) and untuned (Figure 5E) RSFs. A distinction between patients is made based on histological response, which was identified as a strong predictor across all methods (Table 2, Figure 4). For all models, we note that the patient order with respect to histological response was consistent across the Cox, SNN, and all three RSF models.

**Figure 5.**
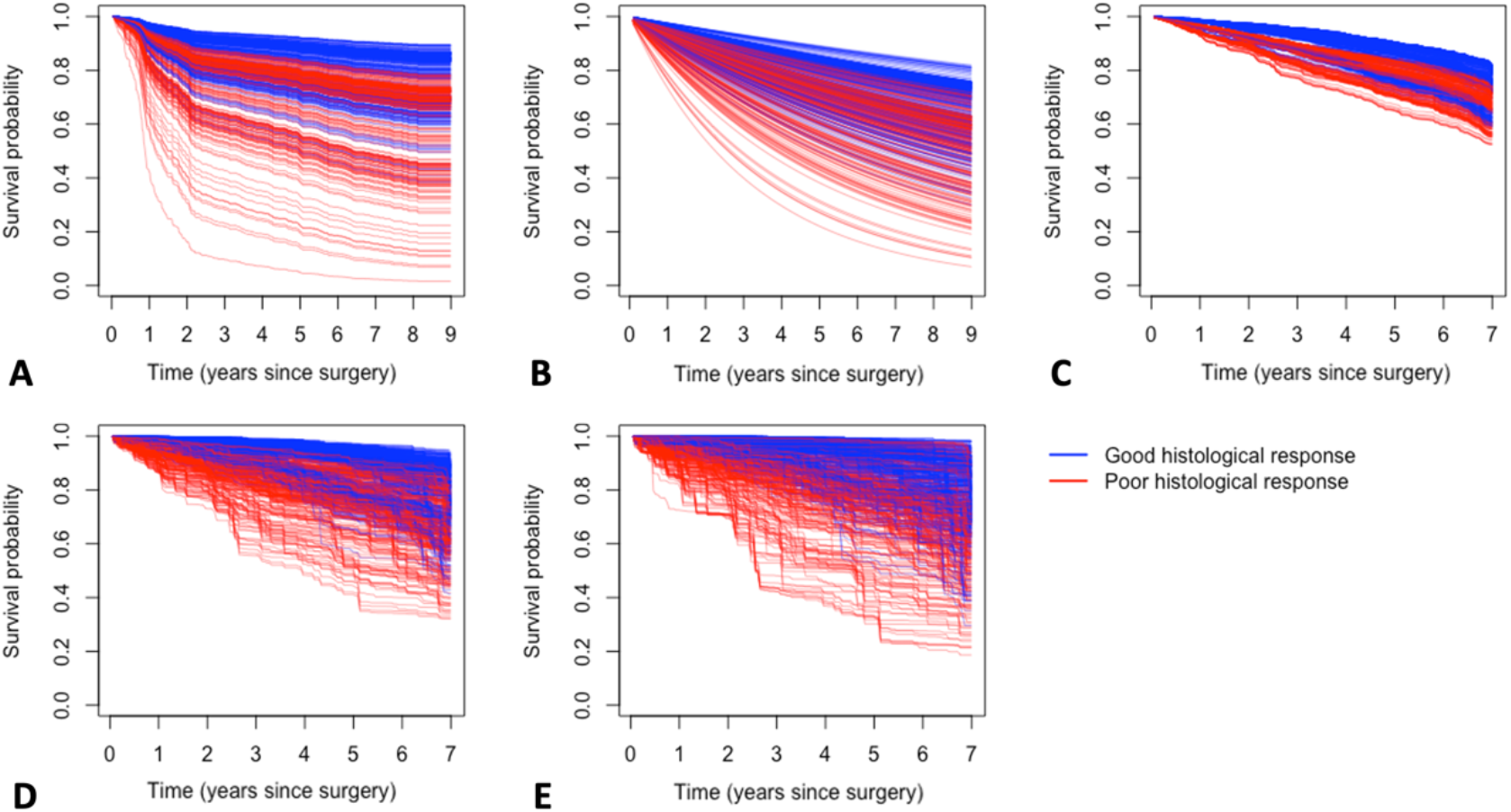
Survival predictions for all validation set patients (N=677), with a distinction made between patients with a poor histological response (red) and a good histological response (blue). Plots are shown for the **A)** Cox proportional hazards model**; B)** SNN; **C)** fully tuned RSF model; **D)** RSF model with *nodesize* and *nodedepth* relaxed; **E)** untuned RSF model with all parameters unrestrained.

In Figure 6, we compare individual survival curves, defining three specific patients with traits of increasing severity, with severity informed by the variable importance values of Figure 4 and Table 2: (i) *reference*: a patient with reference values for all categorical predictors and mean values for the continuous predictors (see also Table 1): female, good histological response, no lung metastases, no other metastases, a radical/wide exicison and mean values for absolute tumour volume and age; (ii) *poor hist*: a patient with reference/mean values for all predictors, but with a poor histological response; *(iii) poor hist + lung mets*: A patient with reference/mean values for all predictors, but with a poor histological response and lung metestases.

**Figure 6.**
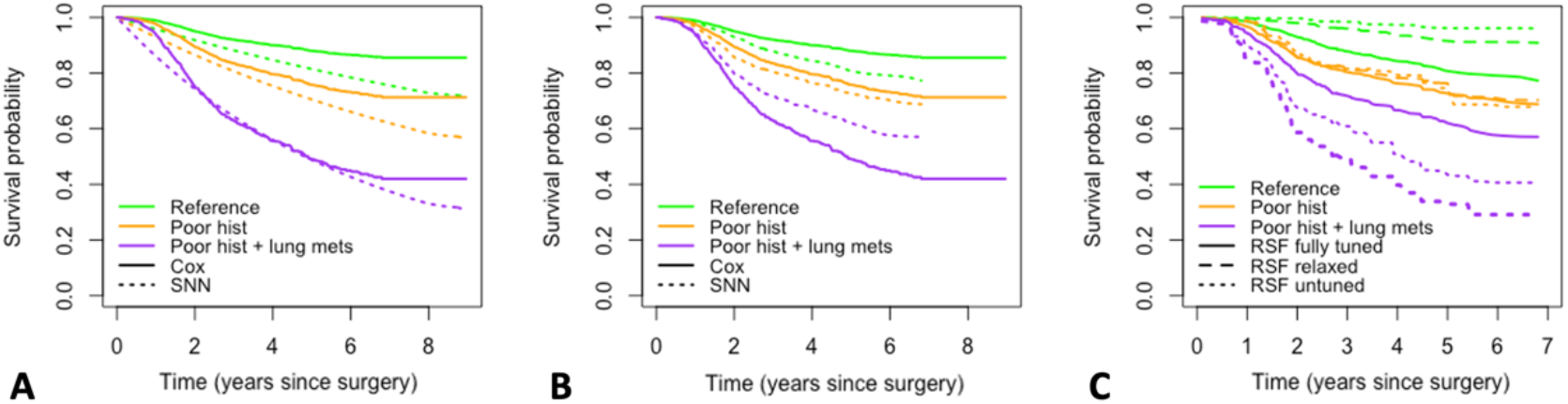
Patient specific predictions for a *reference* patient (green) with reference values for categorical predictors (Table 2) and mean age and volume, a *poor hist* patient (orange) with a poor histological response, and a *poor hist+lung mets* patient (purple), with a poor histological response and lung metastases. **A)** predictions for the Cox proportional hazards model (solid line) and SNN (dotted line); **B)** Predictions for the Cox proportional hazards model (solid line) and fully tuned RSF (dotted line); **C)** predictions for the fully tuned RSF (solid line), the RSF with parameters *nodesize* and *nodedepth* relaxed (dashed line) and the untuned RSF with all parameters unrestrained (dotted line).

All three patient-specific Cox and SNN predictions were similar in the first half of follow-up, with lower SNN predictions in the second half (Figure 6A). The predictions for the fully tuned RSF (Figure 6B) were clustered, with, compared to the Cox predictions, under-and overestimation of the survival probabilities for the *reference* and *poor hist + lung mets* patients, respectively. In contrast, the untuned RSF model over- and underestimated the survival probabilities of these same patients, while the relaxed RSF predictions matched the Cox model most closely.

### 4.5 RSF and SNN instability

ML methods rely on an algorithmic approach, and have intrinsic variability, with repeated runs resulting in variation of the estimates, which, for a well-tuned stable model, will be negligible. Figures 7.1A and 7.2A show the patient-specific survival curves for the fully tuned SNN and RSF models, previously shown in Figures 6A and 6B, on refitting the same model five times. For both, predictions were stable across multiple runs. On relaxing regularization for the SNN, decreased weight decay parameters of 0.015 (Figure 7.1A) and 0.005 (Figure 7.1B) resulted in increasingly erratic predictions. In contrast, for the relaxed RSF (Figure 7.2B) and untuned RSF (Figure 7.2C) the predictions remained stable.

**Figure 7.**
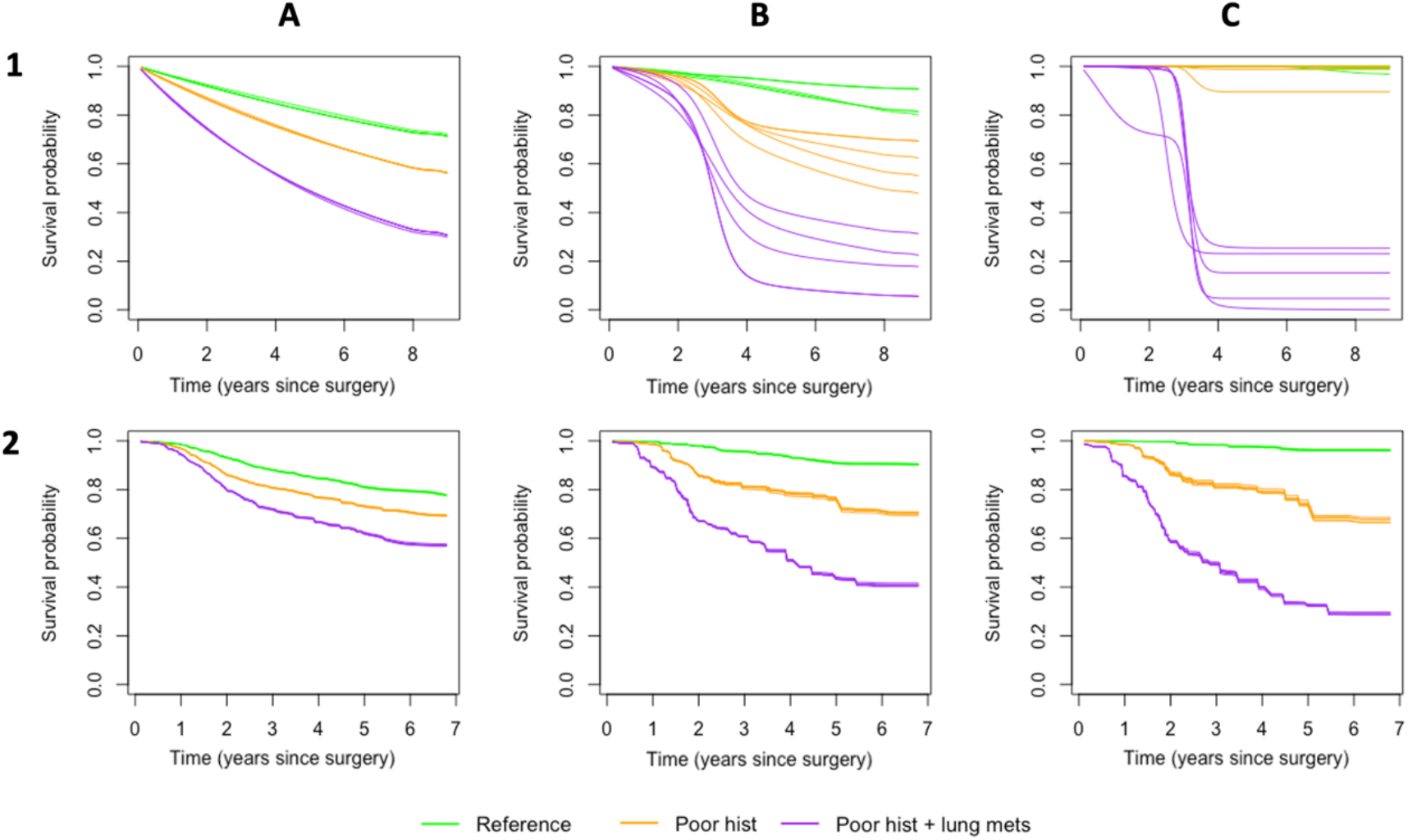
Prediction variability on refitting RSF and SNN models five times. Predictions are shown for a *reference* patient (green) with reference values for categorical predictors (Table 2) and mean age and volume, a *poor hist* patient (orange) with poor histological response, and a *poor hist+lung mets* patient (purple), with poor histological response and lung metastases. **1A)** Prediction curves for five runs of the fully tuned SNN model with 5 hidden nodes and a decay parameter of 0.05; **1B)** Prediction curves for an SNN model with 5 hidden nodes and a decay parameter of 0.015; **1C)** Prediction curves for an SNN model with 5 hidden nodes and a decay parameter of 0.005; **2A)** Prediction curves for the fully tuned RSF model; **2B)** Prediction curves for the RSF model with parameters *nodesize* and *nodedepth* relaxed. **2C)** Prediction curves for the untuned RSF model with all parameters unrestrained

## 5. Discussion

In this paper we applied the Cox proportional hazards model, the survival neural network (SNN), and the random survival forest (RSF) approaches to osteosarcoma data from the EURAMOS-1 clinical trial, and compared them using three performance measures (time-dependent Brier and Kullback-Leibler scores, Harrel’s C-index), the relative variable importance, and patient-specific predictions.

All three methods showed similar performance as measured by the performance measures, with near identical C-indices, and comparable Brier and KL scores. The SNN had slightly lower values in the second half of follow-up (>4.5 years), and predicted lower survival probabilities for individual patients, suggesting that the Cox model may overestimate survival in later follow-up, and that the SNN may have better performance when modelling time periods with sparser events. A limiting factor in the comparison with RSF was the prediction time truncation at 6.9 years. In RSFs, validation set predictions are obtained by ‘dropping’ a patient’s vector down the trees to one of the terminal nodes, which are informed by the event times observed in the training set (Ishwaran *et al*. 2008; Ishwaran & Kogalur, 2008).This precludes extrapolation of predictions to unobserved time points. When interpreting SNN comparisons it should be noted that the model necessitates the division of survival into intervals (Biganzoli *et al*., 1998; Kantidakis *et al*., 2020). We partitioned the follow-up of 9 years in comparatively small, one month-long intervals to facilitate comparison with the Cox model and RSF, which use exact survival times. The effect of interval width has not been sufficiently investigated, and merits consideration, as interval width and the resulting number of observations per interval will impact the variability of the point estimates, their interpretation, and the fit of the neural network itself.

This application highlights an essential unique quality of ML methods, which, unlike statistical models, rely on hyperparameter tuning to identify the best model for a given dataset. This results in within-model variability on refitting a model with the same parametrization, and in-method variability on fitting multiple models of the same method but with different parametrization. As we saw for the fully tuned SNN and RSF, the former is negligible for a well-tuned model. Decreasing regularization for the SNN, however, yielded increasingly erratic individual patient survival curves, and we recommend fitting the same model multiple times to verify the specified hyperparameter values lead to consistent predictions. We observed greater stability for the random survival forest, where foregoing tuning and using default software settings resulted in models with consistent predictions on refitting, and performance measure values comparable to those of the fully tuned RSF. There was, however, considerable in-method variability, with clustered survival curves for the fully tuned RSF, where hyperparameters *nodesize* and *nodedepth* were restricted. This difference in predicted values was not reflected in performance measure values.

The C-index is a measure of discriminative ability and calculates the proportion of pairs of subject that are concordant, with concordance meaning that for a pair of subjects, a comparatively higher risk score (e.g., prognostic index) should be matched by a shorter observed survival time (Harrel *et al*. 1969). The C-index is invariant to scale and given that the relative risk order is maintained, the same value can be obtained under a variety of prediction patterns, and an excellent model performance can be measured, even when the predicted survival curves are unrealistic (e.g., clustered). The time-dependent Bier and KL scores were also near identical for all three RSF models, as were the mean predicted survival curves, indicating that the clustering of predicted survival curves is likely symmetric around the mean. The Brier and KL scores quantify prediction error by comparing the predicted survival of a given patient to the observed survival status at a given time, with a good model predicting low and high survival probabilities for patients with and without events, respectively (Van Houwelingen & Putter, 2012). If the shift of predicted survival probabilities is symmetric around the mean, the prediction error incurred by e.g., overestimating the survival probability of a patient with an event will be proportional to the decrease in prediction error from (correctly) assigning a higher survival probability to a patient with no event, and the total prediction error will, on balance, be the same for all three models.

While the fully tuned RSF model was overall similar to the Cox and SNN models in relative predictor importance, comparatively less importance was accorded to the predictor ‘presence of other metastases’, and more importance to the predictor ‘volume’. Presence of other metastases has in previous literature been identified as a strong predictor for overall survival (Bacci *et al*., 1990; Bielack *et al*., 2002; Rosen *et al*., 1976; Whelan *et al*., 2012), and was found to have high importance for the Cox and SNN models in our application, suggesting that its predictor importance may have been underestimated by the RSF. This can be explained by considering the structure of decision trees, which are grown via binary recursive partitioning. This can be detrimental to predictors with uneven classes, such as this one, where only 79 out of 2032 patients presented with other metastases. Unless an unbalanced predictor is selected as a splitting variable in an early node split, it is unlikely to be selected at all, as each split reduces the number of patients per node, increasing the likelihood that too few or no patients have the relevant characteristic. This is compounded by the presence of strong predictors with more balanced categories, such as ‘histological response’ and ‘presence of lung metastases’. The likelihood of an unbalanced predictor being chosen as a splitting variable is higher for smaller values of *mtry*, and if we compare the tuned RSF (*mtry*=3) to the untuned RSF (*mtry*=7), we note that the relative importance was more than halved for the untuned model. Additionally, we observed that the importance of volume as a predictor was comparatively greater for the untuned RSF, where the greater number of splitting points (*nsplit*=10) favours selection of continuous variables over categorical ones (Ishwaran & Kogalur, 2098).

RSFs are a ML approach that generates consistent survival predictions under the same parameterization but may have very different predictions and variable contributions for different hyperparameter values. RSFs showed a tendency for underestimating the importance of predictors with uneven class distributions, and a preference for selecting continuous predictors when a high number of splitting points was specified. The former may partly be mitigated by restricting the number of candidate variables for node splits. We caution against using RSF for predicting patient-specific survival, as standard model tuning practices may result in aggregated predictions – a quality that is not reflected in the performance measures, as the relative risk ordering remains unchanged. While Cox and RSF model performance can readily be assessed with Harrel’s C-index (Harell *et al*., 1969), with the latter using ensemble mortality as a prognostic index, SNNs require that a time-dependent nonlinear prognostic index be first obtained and subsequently averaged over time. While prognostic index assumes that there exists a natural risk ordering of patients, such monotonicity is not a given in ML models like SNN and RSF, where time-dependent interactions may cause the survival curves to intersect, and the risk order to change (Antolini *et al*., 2005). In our application, we observed high C-indices for both models, indicating excellent discriminative ability. However, if employing SNN or RSF for practices like risk-grouping, we recommend performing a time-dependent assessment prior to applying summary measures. When fitting SNNs, additional factors to consider are the choice of width of the time intervals and the SNN propensity for in-model variability, which should be assessed through repeated model runs.

While ML methods can contribute valuable insights, great care should be taken in their implementation. It is insufficient to assess ML model performance solely on summary performance measures, and any results should be interpreted in context of the specific method’s limitations. In our application to osteosarcoma data from the EURAMOS-1 clinical trial, we investigated the effect of seven predictors on overall survival in 2032 patients, finding little added benefit of RSF and SNN models, when compared to the Cox model. Considering the challenges associated with each method and the loss of straightforward predictor effect interpretation, which is indigenous to ML methods, we recommend using the Cox proportional hazards model for clinical data with a comparatively small number of variables and reserving the use of RSF and SNN for exploratory analyses. A previous simulation study comparing SNNs with the Cox model reached comparable conclusions for clinical trial data, observing that SNNs reached a predictive performance comparable to the Cox model, but were generally less well-calibrated (Kantidakis *et al*., 2021). Part of the results of this study have previously been reported in a master’s thesis (Hazewinkel, 2018).

## Data Availability

Access to the clinical trial data used to support the findings of this study is restricted. Access to the EURAMOS-1 trial data may be requested from the MRC Clinical Trials Unit (CTU, London), who will submit the data application to the Coordinating Data Center (CDC, London), with access subject to review by the Trial Management Group (TMG) and the Trial Steering Committee (TSC).

## Acknowledgements

We thank the EURAMOS-1 trial investigators and participants..

## Ethics Statement

Permission to recruit patients to the protocol was provided by the appropriate national and local regulatory and local committees. Link anonymized data were used for the purposes of this study, and the use of the data was in accordance with the consent taken.

